# DNA methylation age acceleration, depression and anxiety: evidence from a longitudinal adolescent cohort study (SCAMP) and Mendelian Randomisation

**DOI:** 10.64898/2026.09.15.26362972

**Authors:** Chen Shen, Oliver Robinson, Paolo Vineis, Dougal S Hargreaves, Dasha Nicholls, Abbas Dehghan, Mireille B Toledano

## Abstract

**Background:** DNA methylation (DNAm) based age acceleration is an indicator of biological ageing. However, it is unclear how age acceleration is linked to mental health in adolescents. We examined the longitudinal associations between DNAm age acceleration and depression and anxiety in adolescents. We also triangulated the evidence using Mendelian Randomisation (MR).

**Methods:** We analysed a subsample of adolescents (n=261) from a London-based adolescent cohort study. DNAm profile was generated from saliva samples collected at ages 11-12 years. DNAm age acceleration measures were derived from Horvath, PhenoAge, and PedBE clocks. Depressive and anxiety symptoms were measured at ages 13-15 years, using the Patient Health Questionnaire and Generalised Anxiety Disorder scale, respectively. The associations between DNAm age acceleration and depressive and anxiety symptoms were assessed using logistic regression. We also conducted a two-sample MR to investigate the potential causal relationship between Horvath and PhenoAge acceleration and depression and anxiety.

**Results:** We found that PhenoAge age acceleration was associated with higher severity levels of depressive symptoms (OR=1.04, 95% CI 1.01, 1.08) and clinically significant depressive symptoms (OR=1.05, 95% CI 1.01, 1.10). None of these three clocks was associated with anxiety outcomes. MR suggested a potential causal relationship between PhenoAge age acceleration and depression (Inverse variance weighted OR = 1.01, 95% CI 1.00, 1.02).

**Conclusions:** PhenoAge acceleration at ages 11-12 years was associated with depressive symptoms at ages 13-15 years. The association patterns are consistent between longitudinal and MR analyses. PhenoAge acceleration may represent a potential early biomarker for subsequent depressive symptoms in adolescents.

## 1. Introduction

Adolescence is a sensitive period for the onset of mental illness, of which depression and anxiety are the most common manifestations.^1^ In England, more than one in four young people aged 16-24 years had a mental disorder in 2024.^2^ In 2023-2024, ~320,000 young people waited for treatment after being referred to Children and Young People’s Mental Health Services, highlighting the urgent need for early prevention and intervention to reduce this burden.^3^ Depression and anxiety in adolescents have long-lasting negative impacts on their health and personal development.^4 5^

It has been shown that accelerated biological ageing, whereby an individual’s organ systems and tissues age older than their chronological age, predicts the risk of depression and anxiety in adults.^6^ Poor physical health, chronic inflammation, and changes in brain structure and function due to biological ageing are plausible mechanisms.^7–9^ Accelerated ageing in adolescence may also relate to more rapid physical maturation relative to cognitive and emotional development, which is a risk factor for mental health problems.^10 11^ Epigenetic clocks, developed using penalised regression models based on DNA methylation (DNAm) patterns across a set of cytosine-phosphate-guanine (CpG) sites, are among the most widely used molecular estimators of biological age.^12^ For instance, some epigenetic clocks (e.g., Horvath^13^, Hannum^14^, PedBE^15^) were developed to estimate biological age based on age-related CpG sites (clock CpGs), with PedBE specifically developed in paediatric buccal samples. Other clocks (e.g., PhenoAge^16^ and GrimAge^17^) were developed to capture biological ageing by incorporating CpG sites associated with clinical biomarkers, proteins, and ageing-related health outcomes.

Current evidence on the relationship between DNAm age acceleration and mental health in adolescents has been drawn from a mixture of cross-sectional and longitudinal studies. However, cross-sectional studies are subject to reverse causation given the potentially bidirectional relationship between DNAm and mental health.^18 19^ Evidence from longitudinal studies may help clarify temporality. Previous longitudinal studies on DNAm age acceleration and mental health (sample sizes ranging from 148 to 2039) have produced inconsistent findings.^20–24^ Two studies show that age acceleration as estimated by the Horvath and PhenoAge clocks was associated with internalising symptoms.^20 21^ However, two other studies found no association between Horvath age acceleration and internalising symptoms.^22 23^ An association of PedBE, but not Horvath age acceleration, with depressive symptoms has been found in a study.^24^ However, existing studies have largely focused on broad internalising symptom measures rather than employing clinically significant depression or anxiety cut-offs, limiting the clinical applicability of these findings. In addition, causal inference from these studies is limited due to potential residual confounding and reverse causation attributed to the observational study design.

Mendelian Randomisation (MR) is a statistical approach that uses genetic variants associated with the exposure of interest as instrumental variables. Given that genetic alleles are randomly allocated at conception, MR design is analogous to a randomised controlled trial under certain assumptions and is therefore less prone to residual confounding and reverse causation.^25^ The availability of large-scale genome-wide association studies (GWAS) on DNAm age acceleration, depression, and anxiety offers a unique opportunity to provide complementary evidence alongside that from observational studies.

Our study aims to assess the longitudinal associations between DNAm age acceleration and depression and anxiety in adolescents, leveraging a subset of a large-scale London-based adolescent cohort study (the Study of Cognition, Adolescents, and Mobile Phones (SCAMP)). Our study also aims to triangulate evidence from observational findings using two-sample MR to assess whether the association patterns between DNAm age acceleration and depression and anxiety are consistent across complementary study designs.

## 2. Methods

### 2.1 Participants

SCAMP is a school-based longitudinal adolescent cohort study in Greater London, investigating the effects of digital (e.g., mobile phone use, social media use, video gaming) and physical environments (e.g., air pollution, noise, greenspace) on young people’s mental health and cognition.^26^ Eligible schools where participants were recruited were identified using Department of Education school register and 2012 school census.^26^ Participants are representative of the general school-aged children in Greater London. Between November 2014 and July 2016, baseline data were collected from 6590 School Year 7 pupils (aged 11-12 years) across 39 secondary schools. Of them, 3814 pupils from 31 schools participated in the follow-up data collection between November 2016 and July 2018, when they were in School Year 9/10 (aged 13-15 years).

SCAMP pupils participated in a computer-based assessment at baseline and follow-up in examination conditions. The assessment included a battery of cognitive tasks and detailed questionnaires on digital technology use, behaviour, physical and mental health, lifestyle, and socio-demographic characteristics. A subset of SCAMP schools also participated in SCAMP “*Bio-Zone”* at baseline (12 schools) and follow-up (10 schools). Participants from these schools provided non-invasive biological samples and undertook anthropometric measurements. A total of 704 Bio-Zone participants provided saliva samples at baseline and completed follow-up mental health assessments. We selected 264 samples for DNAm profiling, comprising 120 cases (out of 135 total cases) and 144 controls (out of 569 total controls). Cases were defined as participants with clinically significant depressive or anxiety symptoms. Cases and controls were randomly selected within sex strata. This subset was enriched in depression and anxiety cases to maximise the statistical power. The flowchart of the sample selection process is shown in Figure 1.

**Figure 1.**
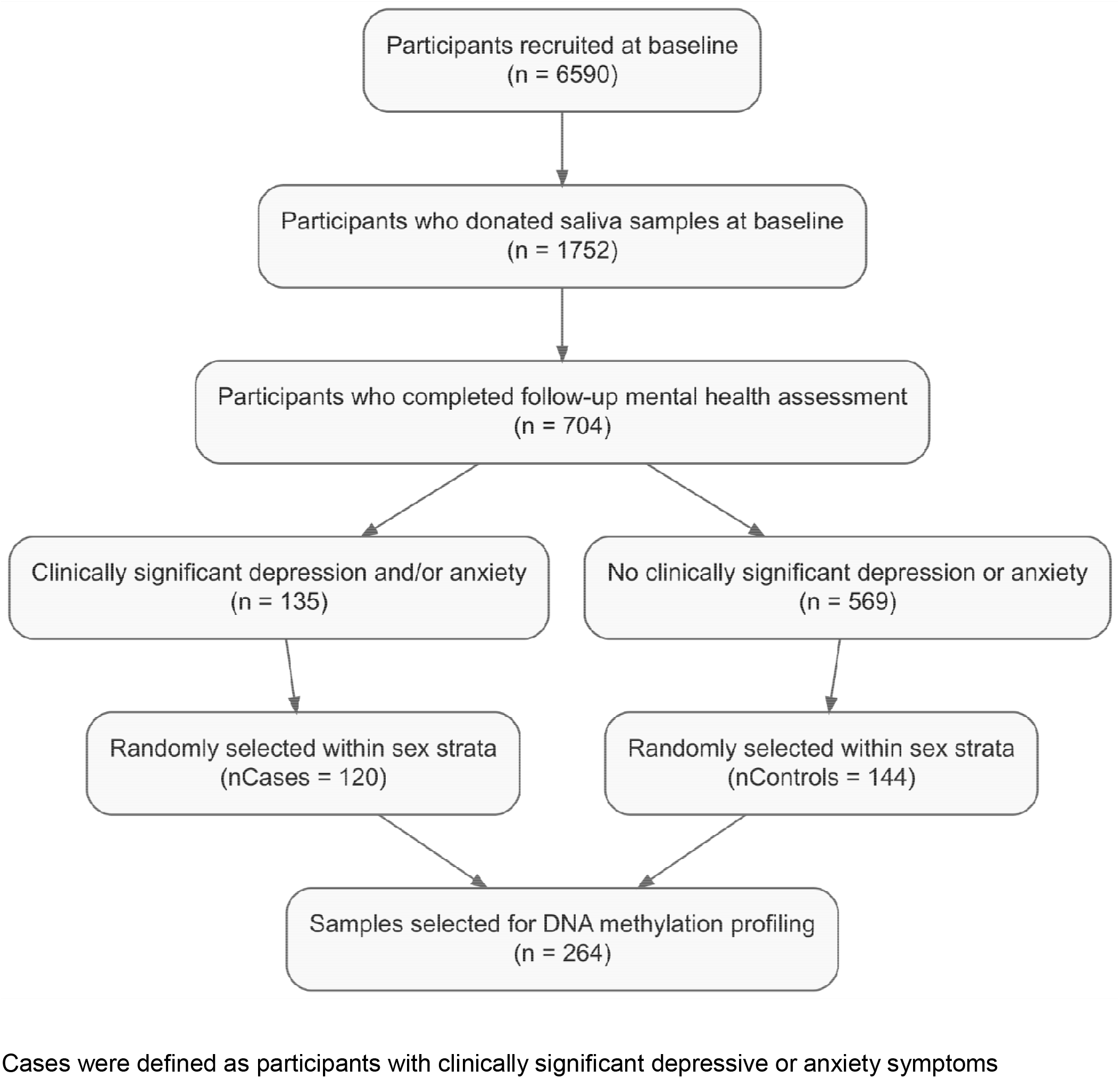
Flowchart for sample selection process for the DNA methylation profiling in the SCAMP cohort. Cases were defined as participants with clinically significant depressive or anxiety symptoms

DNAm data, generated by the Illumina Methylation Screening Array including ~280,000 CpG sites^27^. Quality control procedures were applied to correct dye bias and exclude poor-quality samples and probes.^27^ Samples with a probe detection rate < 0.7 and probes with a call rate < 0.95 were removed. We also removed cross-hybridising and polymorphic probes. A total of 261 participants with 243,887 CpG probes per sample were included in the present analysis.

The SCAMP protocol and subsequent amendments were approved by the North-West Haydock Research Ethics Committee (14/NW/0347). Headteachers of schools consented to participate in SCAMP. Participants were provided in advance with written information about the study and were given the opportunity to opt out of the research at any time. The study was conducted in accordance with the Declaration of Helsinki.

### 2.2 Measures

#### Exposure: DNAm age acceleration at baseline

DNAm age was estimated using established epigenetic clocks. To minimise multiple testing, we selected three clocks (Horvath^13^, PhenoAge^16^, and PedBE^15^) as *a-priori* as they have previously been associated with mental health problems in children and adolescents.^20 21 24^ We used the R package “methylclock” to calculate DNAm age, with the cell-type reference set to “saliva” to adjust for cell-type composition in saliva samples (a mixture of immune, epithelial, and fibroblast cells).^28^ DNAm age acceleration was derived from the residuals of the regression of DNAm age on chronological age.

#### Outcome: depressive and anxiety symptoms at follow-up

Depressive and anxiety symptoms were measured at follow-up. We used the 9-item Patient Health Questionnaire (PHQ-9) to assess depressive symptoms.^29^ A higher PHQ-9 score indicates more depressive symptoms. Depressive symptom severity was categorised as no or minimal (0-4), mild (5-10), moderate (11-14), and moderately severe or severe (15+). A summary score of 11 or higher indicates clinically significant depressive symptoms. We used the 7-item Generalised Anxiety Disorder (GAD-7) scale to assess anxiety symptoms.^30^ A higher GAD-7 score indicates more anxiety symptoms. Anxiety symptom severity was categorised as no or minimal (0-4), mild (5-9), moderate (10-14), and moderately severe or severe (15+). A summary score of 10 or higher indicates clinically significant anxiety symptoms. Both PHQ-9 and GAD-7 have demonstrated good sensitivity and specificity for detecting clinically significant symptoms in adolescents.^29 31^

#### Covariates

Covariates included age at follow-up, gender, ethnicity, parental socioeconomic status (SES), area-level deprivation, school type (state versus independent), and baseline body mass index (BMI) z-score and emotional difficulties. We categorised ethnicity as White, Black, Asian, and Mixed/others. Parental SES was ascertained from self-reported parental occupation and then converted to three levels (managerial and professional, intermediate, and routine or manual) based on the National Statistics Socio-economic classification (NS-SEC). Participants were asked to report both their father’s and mother’s occupations. Participants were assigned the higher NS-SEC level if the NS-SEC levels differed between parents. Area-level deprivation was evaluated using the 2015 Index of Multiple Deprivation (IMD), derived from the postcodes of participants’ residential addresses. IMD is a composite index of deprivation based on weighted scores across seven domains, including income, employment, education, health, crime, housing and service access, and living environment. BMI was derived from measured height and weight in SCAMP Bio-Zone. We derived an age- and sex-specific BMI z-score based on the 2007 WHO growth reference (aged 5-19 years) for each participant.^32^ Emotional difficulties were assessed using the score of the Strengths and Difficulties Questionnaire (SDQ) emotional subscale.^33^

### 2.3 Two-sample MR analysis

We leveraged GWAS summary statistics of DNAm age acceleration (Horvath and PhenoAge clocks), depression, and anxiety.^34–36^ GWAS for PedBE age acceleration is currently unavailable. GWAS for DNAm age acceleration includes 34,710 participants. GWAS for depression includes 525,197 cases and 3,362,335 controls. GWAS for anxiety includes 122,341 cases and 729,881 controls. Depression and anxiety cases from these GWAS were ascertained based on multiple methods, e.g., clinical interview, questionnaire, and self-reported diagnosis. The genomic structural equation modelling analysis indicated that depression and anxiety phenotypes ascertained by different methods had largely consistent underlying genetic liability. We therefore used the GWAS summary statistics from the full samples to maximise statistical power of MR. We only performed MR in European ancestry due to data availability.

We selected the single-nucleotide polymorphisms (SNPs) associated with DNAm age acceleration at genome-wide significance (P < 5×10^−8^). We clumped the SNPs for linkage disequilibrium using the 1000 Genomes Project reference panel based on a threshold of R^2^ < 0.001 and a clumping distance of 250 kb. We also calculated the F-statistics to assess instrument strength, with F-statistics < 10 indicating weak instrument bias.^37^

### 2.4 Statistical analysis

#### 2.4.1 Observational study in SCAMP

We used multivariable ordinal logistic regression to assess the associations between DNAm age acceleration at baseline and depressive and anxiety symptom severity at follow-up. We used multivariable logistic regression to assess the associations between DNAm age acceleration at baseline and clinically significant depressive and anxiety symptoms at follow-up. Potential confounders included age at follow-up, gender, ethnicity, parental SES, IMD, school type, baseline score of SDQ emotional subscale and baseline BMI z-score, selected *a priori*. Given the small sample size (<10) from several schools, we did not control for the school clustering effect as this would produce unstable estimates. We also examined whether these associations varied by gender (male and female) or ethnicity (white and non-white) by testing the significance of the interaction term.

We used multiple imputation to predict missing data on baseline emotional difficulties (6.5%) and BMI z-score (15.7%) using a flexible additive regression model. In the imputation model, we included DNAm age acceleration estimates, PHQ-9 and GAD-7 scores, gender, age at follow-up, parental SES, IMD score, school type, and auxiliary variables including scores of SDQ subscales and wellbeing (measured by KIDSCREEN-10^38^) at follow-up. We generated 50 imputed datasets and yielded pooled effect estimates and 95% CIs that incorporated uncertainty due to missing data.

We performed sensitivity analyses to assess the robustness of the observational findings. First, we excluded participants with baseline emotional difficulties (the score of SDQ emotional subscale ≥ 6) to assess the associations between DNAm age acceleration and depressive and anxiety symptoms in participants without pre-existing emotional difficulties. Second, we used complete case analysis to assess the associations between DNAm age acceleration and depressive and anxiety symptoms in those with complete data on baseline emotional difficulties and BMI.

#### 2.4.2 MR study

The random-effect inverse variance weighted (IVW) method^39^ was used as the primary MR approach to estimate the effects of DNAm age acceleration on depression and anxiety. We also used the weighted median^40^ to assess the robustness of the results. We used MR Egger^41^ to assess potential horizontal pleiotropy (i.e., that the effect of the genetic instrument on depression or anxiety is through an alternative pathway rather than DNAm age acceleration) based on the significance of the Egger intercept term. The heterogeneity of genetic instruments was assessed using Cochran’s Q statistic.

We also performed some MR sensitivity analyses. Specifically, we used a leave-one-out analysis to evaluate whether the causal effect was driven by a single instrument. We used MR-Pleiotropy RESidual Sum and Outlier (MR-PRESSO)^42^ analysis to detect potential outlier instruments. We used the MR Steiger test^43^ to further explore the likely direction of causality between DNAm age acceleration and depression or anxiety. All analyses were performed using R (version 4.4.2).

## 3. Results

### 3.1 Findings from the SCAMP cohort study

Table 1 shows the descriptive statistics of the subset of the SCAMP cohort included in this study. The sample was enriched for participants with clinically significant depressive or anxiety symptoms, with prevalences of 33.3% and 31.4%, respectively. The mean age was 11.76 years at baseline and 13.99 years at follow-up. About two-thirds of participants were female (63.2%). The sample was diverse in terms of ethnicity and parental SES. Table S1 shows the descriptive statistics in the case and control samples.

**Table 1.** Descriptive statistics of the participants (n=261)

|  |  |
| --- | --- |
| **Demographic variables** |  |
| Age (years) at baseline, mean (SD) | 11.76 (0.43) |
| Age (years) at follow-up, mean (SD) | 13.99 (0.52) |
| Gender, n (%) |  |
| Male | 96 (36.8) |
| Female | 165 (63.2) |
| Ethnicity, n (%) |  |
| White | 108 (41.4) |
| Black | 39 (14.9) |
| Asian | 93 (35.6) |
| Mixed/others | 21 (8.1) |
| Parental socioeconomic status, n (%) |  |
| Managerial/professional occupations | 147 (56.3) |
| Intermediate occupations | 65 (24.9) |
| Routine and manual occupations | 49 (18.8) |
| Index of deprivation (quintile) |  |
| 1 (most deprived) | 75 (28.7) |
| 2 | 73 (28.0) |
| 3 | 53 (20.3) |
| 4 | 40 (15.3) |
| 5 (least deprived) | 20 (7.7) |
| Type of school, n (%) |  |
| Independent | 67 (25.7) |
| State | 194 (74.3) |
| **Clock-estimated DNA methylation age (baseline)** |  |
| Horvath | 13.22 (2.55) |
| PhenoAge | 13.75 (6.78) |
| PedBE | 8.48 (0.95) |
| **Outcomes (follow-up)** |  |
| Depression severity, n (%) |  |
| No/minimal | 104 (39.9) |
| Mild | 70 (26.8) |
| Moderate | 41 (15.7) |
| Severe | 46 (17.6) |
| Anxiety severity, n (%) |  |
| No/minimal | 111 (42.5) |
| Mild | 68 (26.1) |
| Moderate | 53 (20.3) |
| Severe | 29 (11.1) |
| Presence of clinical depression, n (%) | 87/261 (33.3) |
| Presence of clinical anxiety, n (%) | 82/261 (31.4) |

Table 2 shows that after adjusting for age at follow-up, sex, ethnicity, parental SES, area-level deprivation, school type, and baseline emotional difficulties, per year increase in PhenoAge acceleration at baseline was associated with increased depressive symptom severity and higher odds of clinically significant depressive symptoms at follow-up. The associations remained similar after additional adjustment for baseline BMI z-score (symptom severity OR = 1.04, 95% CI 1.01, 1.08; clinically significant symptom OR = 1.05, 95% CI 1.01, 1.10). The associations between PhenoAge acceleration and depression outcomes did not vary by sex or ethnicity (both P values for interaction > 0.05). None of the three DNAm age acceleration measures was associated with anxiety outcomes.

**Table 2.** Associations between DNA methylation age acceleration at baseline and depression and anxiety at follow-up (n=261)

| Mental health measures | Clocks | Unadjusted model<br>OR (95% CI) | Model 1<br>OR (95% CI) | Model 2<br>OR (95% CI) |
| --- | --- | --- | --- | --- |
| Depression severity | Horvath | 1.06 (0.97, 1.16) | 1.10 (1.00, 1.21) | 1.09 (0.99, 1.20) |
|  | PhenoAge | 1.04 (1.01, 1.07) | 1.05 (1.01, 1.08) | 1.04 (1.01, 1.08) |
|  | PedBE | 0.93 (0.73, 1.17) | 0.98 (0.75, 1.28) | 0.98 (0.75, 1.28) |
| Anxiety severity | Horvath | 1.04 (0.95, 1.14) | 1.05 (0.96, 1.16) | 1.04 (0.94, 1.14) |
|  | PhenoAge | 1.03 (1.00, 1.07) | 1.03 (0.99, 1.07) | 1.02 (0.99, 1.06) |
|  | PedBE | 0.84 (0.66, 1.06) | 0.84 (0.64, 1.11) | 0.83 (0.63, 1.10) |
| Clinical depression | Horvath | 1.07 (0.97, 1.18) | 1.10 (0.98, 1.23) | 1.10 (0.98, 1.23) |
|  | PhenoAge | 1.05 (1.01, 1.09) | 1.05 (1.01, 1.10) | 1.05 (1.01, 1.10) |
|  | PedBE | 0.85 (0.65, 1.13) | 0.91 (0.66, 1.27) | 0.91 (0.66, 1.27) |
| Clinical anxiety | Horvath | 1.06 (0.96, 1.18) | 1.09 (0.96, 1.22) | 1.07 (0.95, 1.21) |
|  | PhenoAge | 1.03 (0.99, 1.07) | 1.02 (0.97, 1.06) | 1.01 (0.97, 1.06) |
|  | PedBE | 0.95 (0.72, 1.26) | 0.99 (0.70, 1.39) | 1.00 (0.71, 1.40) |

Model 1: Adjusted for age at follow-up, sex, ethnicity, parental socioeconomic status, area-level deprivation, school type, and emotional difficulties at baseline measured by Strengths and Difficulties Questionnaire.
Model 2: Additionally adjusted for BMI z-score at baseline.
Multiple imputation was used to impute missing data on emotional difficulties (6.5%) and BMI z-score (15.7%).

Sensitivity analyses show that the associations between PhenoAge acceleration and depression outcomes remained evident after excluding participants with baseline emotional difficulties (Table S2). The overall association pattern also remained similar, despite variations in effect estimates. Complete case analysis did not alter the pattern, although the associations between PhenoAge acceleration and depression outcomes did not reach the significance level (Table S3).

### 3.2 Findings from the two-sample MR

The number of genetic instruments included in our MR ranged from 11 to 31, with the minimum F-statistic of these instruments ranging from 30.2 to 31.8 (Table S4). Figure 2 provides suggestive evidence of a potential causal relationship between PhenoAge acceleration and depression (IVW OR = 1.01, 95% CI 1.00, 1.02, P = 0.048). The heterogeneity of genetic instruments was not significant but suggestive (P_Q-statistic_ = 0.052, Table S4). The effect estimate was consistent with the weighted median method (OR = 1.01, 95% CI 1.00, 1.02, P = 0.006). No association was observed between PhenoAge acceleration and anxiety, or between Horvath age acceleration and either depression or anxiety.

**Figure 2.**
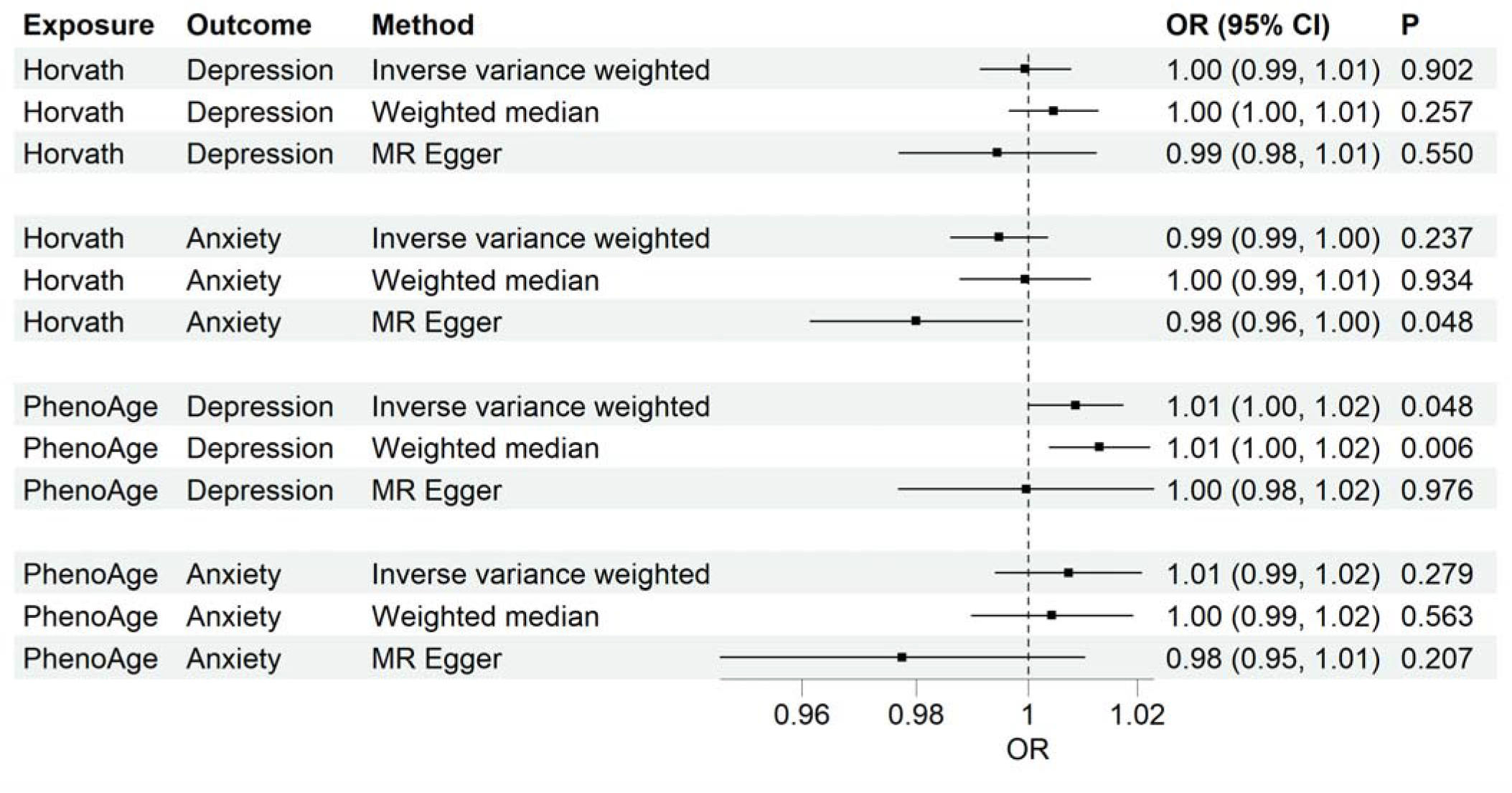
The two-sample Mendelian Randomisation results on DNA methylation age acceleration (Horvath and PhenoAge clocks) and depression and anxiety

MR Egger results indicate no statistical evidence of horizontal pleiotropy (all P-values for the Egger intercept > 0.05, Table S4). Sensitivity analyses showed that the direction of observed associations between DNAm age acceleration and depression and anxiety remained consistent after leaving each individual SNP, except for the association between Horvath age acceleration and depression (Figure S1). MR-PRESSO analysis only detected outlier instruments for the association between Horvath age acceleration and depression (outlier-corrected OR = 1.00, P = 0.836). The MR Steiger test suggests no evidence of reverse causation for any association (all P values < 0.001, Table S4).

## Discussion

Our study combined a longitudinal cohort study and MR to assess the associations between DNAm age acceleration and depression and anxiety. In a longitudinal cohort, we found that PhenoAge acceleration at ages 11-12 years, but not Horvath or PedBE age acceleration, was associated with depressive symptoms at ages 13-15 years. None of these three clocks was associated with anxiety symptoms. Such an association pattern was consistent with MR results.

Our study adds to the evidence base that in adolescents, PhenoAge acceleration was associated with later depressive symptoms, but not anxiety. One observational study (n=2039) did not find any associations between PhenoAge acceleration and depressive and anxiety symptoms at age 15.^20^ This suggests that phenotypic ageing starting at an earlier stage may have a more prominent effect on depressive symptoms. Previous observational studies on age acceleration in relation to depressive or anxiety symptoms produced mixed results. One study (n=71) found that Horvath age acceleration was significantly associated with more severe depressive and anxiety symptoms.^44^ However, this study was conducted in participants who experienced maltreatment, so the results may not be generalisable to the general adolescent population. Another study in a community-based cohort (n=239) also reported an association between Horvath age acceleration and anxious and depressed problems, but their symptoms were mother-reported.^45^ Our findings are consistent with two longitudinal studies that did not find any association of Horvath or PedBE age acceleration with depressive or anxiety symptoms,^22 23^ but another longitudinal study (n=149) found that PedBE age acceleration was associated with the top 30% depressive symptom scores.^24^ However, the top 30% dichotomisation is not based on a clinical cut-off.

Previous MR studies did not find any causal relationship between PhenoAge acceleration and depression or anxiety.^19 46^ Although these two studies used the same GWAS summary statistics for DNAm age acceleration as the present study, the GWAS summary statistics for depression and anxiety were derived from smaller samples, which may have limited statistical power to detect significant associations.

DNAm age acceleration during adolescence may reflect differences in the pace of physical and biological development. Previous studies have found associations between DNAm age acceleration and obesity, whilst obesity is associated with mental health problems.^11 47^ However, additional adjustment for age-and sex-standardised BMI had little impact on the observed associations in our study, suggesting that differences in body size do not explain the findings. In addition to chronological age, PhenoAge clock was derived by incorporating information from clinical biomarkers including the inflammation marker C-reactive protein (CRP).^16^ Although these ageing-related biomarkers may show less variation in children and adolescents than in adults, evidence supports that systemic inflammation may contribute to depression during adolescence. A meta-analysis in children and adolescents has found an association between CRP level and depression.^48^ However, inflammatory biomarkers were not measured in SCAMP participants. Therefore, the role of the inflammatory process linking PhenoAge acceleration and depressive symptoms requires further investigation.

Strengths of our study include that we triangulated evidence using two complementary study designs and yielded consistent findings. The longitudinal study design of the SCAMP cohort allows us to assess the temporal sequence of DNAm age acceleration and subsequent depressive and anxiety symptoms. Our MR Steiger test also confirmed the directionality of the association.

Our study has several limitations. First, the sample size of this DNAm subset is small, and the participants have a narrow age range (11-12 years). However, age acceleration still informs adolescents’ mental health reflected by the underlying biological processes. Second, attrition occurred during follow-up. However, loss to follow-up was largely attributable to factors affecting school participation in follow-up assessments, such as logistical constraints related to school timetables, rather than mental health status of individual participants. Third, we cannot rule out the possibility of residual confounding. For instance, psychosocial stress and pubertal timing may influence both DNAm age acceleration and mental health in adolescents and could partly explain the observed associations.^49 50^ Fourth, the observational findings were based on DNAm from saliva samples in an adolescent cohort, whilst MR findings were based on DNAm from blood samples and predominantly in adult populations. Although DNAm profile varies by age and tissue, previous evidence has shown that the effects of genetic influences on DNAm are largely consistent across tissues and over the lifespan.^51 52^ This supports comparability between observational and MR findings. Fifth, there is partial sample overlap between the exposure and outcome GWAS summary statistics, which may introduce weak instrument bias and inflate MR estimates towards the confounded estimates from observational studies. However, all genetic instruments included in the MR are strong (all F-statistics > 30), indicating that weak instrument bias due to sample overlap is not a major concern. Sixth, GWAS on PedBE age acceleration is not available, so we are not able to conduct the MR to assess its causal relationship with depression and anxiety.

The longitudinal association between PhenoAge acceleration and subsequent depressive symptoms in adolescents suggests that PhenoAge acceleration may represent a promising biomarker associated with future depression risk. Future studies with a larger sample size and participants from different age groups are warranted to replicate our findings. The non-invasive nature of saliva sample collection makes it particularly practicable for large-scale population research about DNAm in school and community settings. Our findings may inform future research into the early identification and prevention of depression. Future studies are warranted to investigate the biological mechanisms underlying the association between PhenoAge acceleration and depression and to determine whether these findings have implications for preventive strategies.

## Conclusions

In an adolescent cohort, PhenoAge acceleration at ages 11-12 years was associated with depressive symptoms at ages 13-15 years. Our MR analysis supports a potential causal relationship between PhenoAge acceleration and depression. The association patterns are consistent between longitudinal and MR analyses. Accelerated epigenetic ageing (estimated by the PhenoAge clock) may represent a potential early biomarker for subsequent depressive symptoms in adolescents.

## Data Availability

According to the participant consent, access to individual-level SCAMP data needs to be approved by the SCAMP Data Access Committee (https://scampstudy.org/get-involved/opportunities-for-researchers/). Application should be made to. The data dictionary is available on request to the corresponding author. The GWAS summary statistics used in this study are publicly available at the following repositories: https://figshare.com/articles/dataset/GWAS_summary_statistics_for_major_depression_PGC_MDD2025_/27061255; https://figshare.com/articles/dataset/anx2026/31389910; https://datashare.ed.ac.uk/items/57cb0570-984b-41da-9fb5-b6831b327487.

## CRediT authorship contribution statement

**Chen Shen**: Writing - review & editing, Writing - original draft, Visualization, Project administration, Methodology, Funding acquisition, Formal analysis, Data curation, Conceptualization. **Oliver Robinson**: Writing - review & editing, Validation, Methodology. **Paolo Vineis**: Writing - review & editing, Methodology, Funding acquisition. **Dougal S Hargreaves**: Writing - review & editing, Methodology, Funding acquisition. **Dasha Nicholls**: Writing - review & editing, Methodology, Funding acquisition. **Abbas Dehghan**: Writing - review & editing, Validation, Methodology. **Mireille B. Toledano**: Writing - review & editing, Supervision, Resources, Project administration, Investigation, Funding acquisition.

