## Supplementary tables and figures for "DNA methylation age acceleration, depression and anxiety: evidence from a longitudinal adolescent cohort study (SCAMP) and Mendelian Randomisation"

Table S1 Descriptive statistics in case and control samples

| **Demographic variables** | Depression  (n=87) | Anxiety  (n=82) | Control (n=142) |
| --- | --- | --- | --- |
| Age (years) at baseline, mean (SD) | 11.71 (0.46) | 11.74 (0.45) | 11.80 (0.41) |
| Age (years) at follow-up, mean (SD) | 14.03 (0.56) | 14.05 (0.58) | 13.96 (0.48) |
| Gender, n (%) |  |  |  |
| Male | 21 (24.1) | 15 (18.3) | 68 (47.9) |
| Female | 66 (75.9) | 67 (81.7) | 74 (52.1) |
| Ethnicity, n (%) |  |  |  |
| White | 34 (39.1) | 39 (47.6) | 55 (38.7) |
| Black | 16 (18.4) | 14 (17.1) | 20 (14.1) |
| Asian | 32 (36.8) | 22 (26.8) | 54 (38.0) |
| Mixed/others | 5 (5.8) | 7 (8.5) | 13 (9.2) |
| Parental socioeconomic status^a^, n (%) |  |  |  |
| Managerial/professional occupations | 51 (58.6) | 50 (61.0) | 77 (54.2) |
| Intermediate occupations | 20 (23.0) | 20 (24.4) | 38 (26.8) |
| Routine and manual occupations | 16 (18.4) | 12 (14.6) | 27 (19.0) |
| Index of deprivation (quintile) |  |  |  |
| 1 (most deprived) | 23 (26.4) | 23 (28.1) | 42 (29.6) |
| 2 | 32 (36.8) | 20 (24.4) | 36 (25.4) |
| 3 | 12 (13.8) | 13 (15.9) | 36 (25.4) |
| 4 | 13 (14.9) | 17 (20.7) | 19 (13.4) |
| 5 (least deprived) | 7 (8.1) | 9 (11.0) | 9 (6.3) |
| Type of school, n (%) |  |  |  |
| Independent | 22 (25.3) | 24 (29.3) | 35 (24.7) |
| State | 65 (74.7) | 58 (70.7) | 107 (75.4) |
| **Clock-estimated DNA methylation age (baseline)** |  |  |  |
| Horvath | 13.53 (2.53) | 13.50 (2.47) | 13.04 (2.64) |
| PhenoAge | 15.23 (7.56) | 14.60 (7.08) | 12.66 (6.20) |
| PedBE | 8.41 (1.03) | 8.46 (0.87) | 8.52 (0.93) |

Depression or anxiety cases were defined as participants with clinically significant depressive or anxiety symptoms

Table S2 Associations between DNA methylation age acceleration at baseline and depression and anxiety at follow-up, excluding participants with emotional difficulties at baseline (n=206)

| Mental health measures | Clocks | OR (95% CI) |
| --- | --- | --- |
| Depression severity | Horvath | 1.05 (0.94, 1.17) |
|  | PhenoAge | 1.04 (1.00, 1.08) |
|  | PedBE | 1.01 (0.75, 1.37) |
| Anxiety severity | Horvath | 0.99 (0.89, 1.11) |
|  | PhenoAge | 1.03 (0.99, 1.07) |
|  | PedBE | 0.84 (0.62, 1.15) |
| Clinical depression | Horvath | 1.08 (0.95, 1.23) |
|  | PhenoAge | 1.05 (1.00, 1.10) |
|  | PedBE | 0.83 (0.57, 1.20) |
| Clinical anxiety | Horvath | 1.04 (0.90, 1.20) |
|  | PhenoAge | 1.01 (0.96, 1.06) |
|  | PedBE | 1.01 (0.69, 1.50) |

Adjusted for age at follow-up, sex, ethnicity, parental socioeconomic status, area-level deprivation, school type, baseline emotional difficulties, and baseline BMI z-score

Table S3 Associations between DNA methylation age acceleration at baseline and depression and anxiety at follow-up using complete-case analysis (n=204)

| Mental health measures | Clocks | OR (95% CI) |
| --- | --- | --- |
| Depression severity | Horvath | 1.05 (0.94, 1.18) |
|  | PhenoAge | 1.04 (0.99, 1.08) |
|  | PedBE | 1.09 (0.80, 1.49) |
| Anxiety severity | Horvath | 1.01 (0.90, 1.13) |
|  | PhenoAge | 1.02 (0.98, 1.07) |
|  | PedBE | 0.86 (0.62, 1.17) |
| Clinical depression | Horvath | 1.10 (0.96, 1.25) |
|  | PhenoAge | 1.05 (1.00, 1.10) |
|  | PedBE | 1.05 (0.71, 1.54) |
| Clinical anxiety | Horvath | 1.05 (0.91, 1.21) |
|  | PhenoAge | 0.99 (0.94, 1.04) |
|  | PedBE | 0.87 (0.58, 1.29) |

Adjusted for age at follow-up, sex, ethnicity, parental socioeconomic status, area-level deprivation, school type, baseline emotional difficulties, and baseline BMI z-score

Table S4 Quality control measures of the two-sample Mendelian Randomisation

| Exposure | Outcome | N_SNP_ | Min F statistics | MR Egger intercept | Egger intercept P value | Q | Q statistics P value | MR Steiger P value |
| --- | --- | --- | --- | --- | --- | --- | --- | --- |
| Horvath | Depression | 27 | 30.2 | 0.002 | 0.542 | 78.4 | <0.001 | <0.001 |
| Horvath | Anxiety | 31 | 30.2 | 0.005 | 0.099 | 38.5 | 0.137 | <0.001 |
| PhenoAge | Depression | 11 | 31.8 | 0.003 | 0.432 | 18.2 | 0.052 | <0.001 |
| PhenoAge | Anxiety | 12 | 31.8 | 0.011 | 0.085 | 17.7 | 0.088 | <0.001 |

N_SNP_: Number of genetic instruments included in each MR.

Figure S1 Mendelian Randomisation leave-one-out analysis for DNA methylation acceleration and depression and anxiety

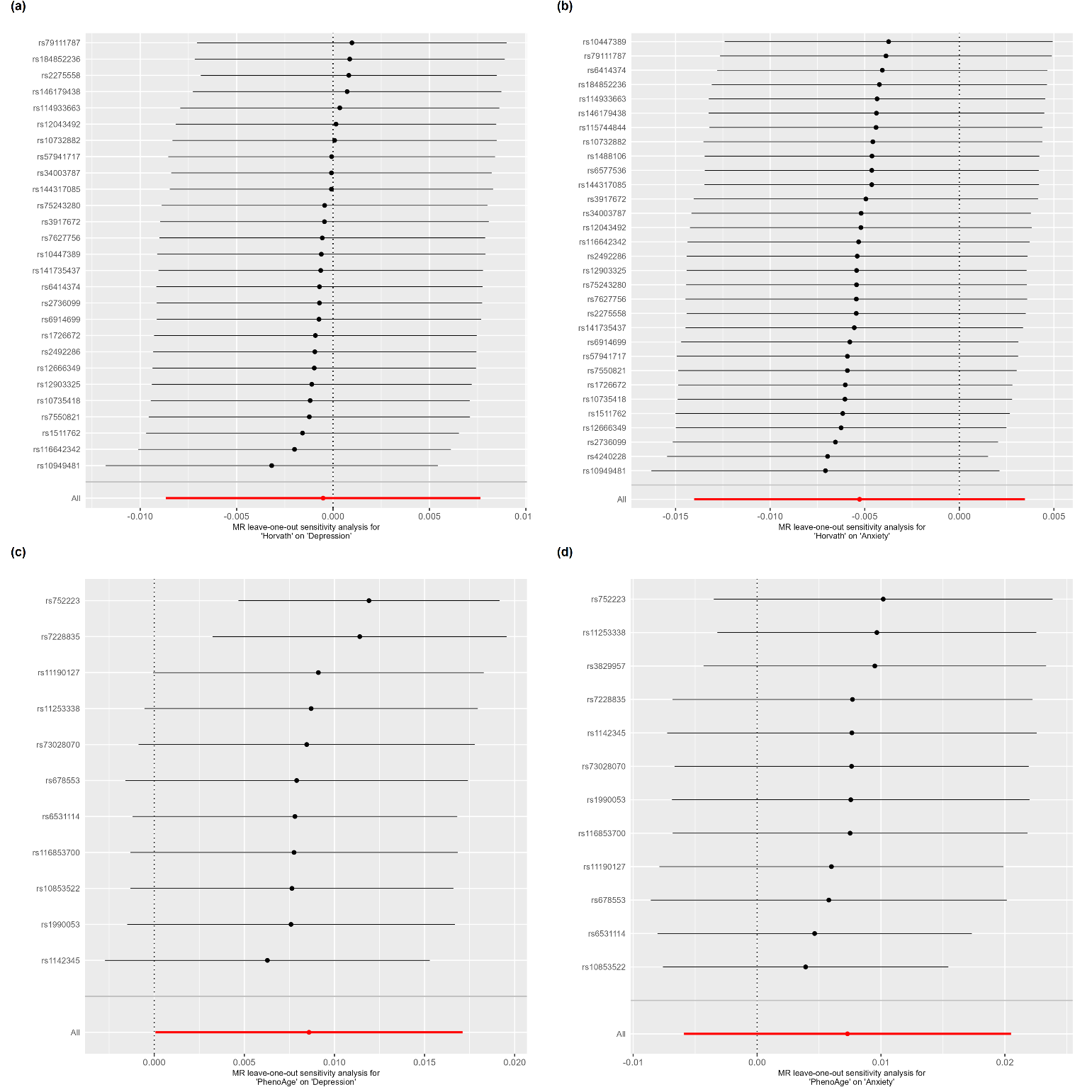

a: Leave-one-out analysis for Horvath age acceleration and depression

b: Leave-one-out analysis for Horvath age acceleration and anxiety

c: Leave-one-out analysis for PhenoAge acceleration and depression

d: Leave-one-out analysis for PhenoAge acceleration and anxiety
